# Photon Absorption Remote Sensing Imaging of Breast Needle Core Biopsies is Diagnostically Equivalent to Gold Standard H&E Histologic Assessment

**DOI:** 10.1101/2023.08.05.23293672

**Authors:** James E.D. Tweel, Benjamin R. Ecclestone, Hager Gaouda, Deepak Dinakaran, Michael P. Wallace, Gilbert Bigras, John R. Mackey, Parsin Haji Reza

## Abstract

**OBJECTIVE:** Photon absorption remote sensing (PARS) is a new laser-based microscope technique that permits cellular level resolution of unstained fresh, frozen, and fixed tissues. Our objective was to determine whether PARS could provide image quality sufficient for diagnostic assessment of breast cancer needle core biopsies (NCB).

**DESIGN:** We PARS imaged and virtually H&E stained seven independent unstained formalin fixed paraffin-embedded breast NCB sections. These identical tissue sections were then subsequently stained with standard H&E and digitally scanned. Both the 40x PARS and H&E whole slide images were assessed by seven breast cancer pathologists, masked to the origin of the images. A concordance analysis was performed to quantify the diagnostic performances of standard H&E and PARS virtual H&E.

**RESULTS:** The PARS images were deemed of diagnostic quality and pathologists were unable to distinguish the origin of the images above that expected by chance. The diagnostic concordance on cancer vs. benign was high between PARS and conventional H&E (98% agreement) and there was complete agreement for within PARS images. Similarly, agreement was substantial (kappa > 0.6) for diagnosis of specific cancer subtypes. PARS virtual H&E inter-rater reliability was broadly consistent with the published literature on diagnostic performance of conventional histology NCBs across all tested histologic features.

**CONCLUSIONS AND RELEVANCE:** PARS was able to provide images on unstained tissues slides that were diagnostically equivalent to conventional H&E. Due to its ability to non-destructively image fixed and fresh tissues, and the suitability of the PARS output for artificial intelligence assistance in diagnosis, this technology has the potential to improve both the speed and accuracy of breast cancer diagnosis.

## 1. INTRODUCTION

A breast needle core biopsy (NCB) is a medical procedure in which a small, cylindrical piece of breast tissue is removed for examination and diagnosis, typically with the aid of imaging guidance (e.g., ultrasound)^1^. The procedure is performed when an abnormality is found in the breast, such as a palpable mass, or an area of suspicious tissue seen on a mammogram or other imaging tests. It is an established standard of care for obtaining accurate preoperative histological diagnosis of suspicious breast lesions^2–7^. In addition, it offers numerous advantages, including reduced cost and complication rates, over surgical biopsies primarily due its minimally invasive approach^8–11^.

Following the NCB procedure, the samples undergo standard tissue processing and staining procedures to enable histological analysis. Samples are formalin fixed and subsequently embedded in paraffin wax where they are thinly sectioned (∼5μm) and placed on glass slides for staining with hematoxylin and eosin (H&E)^12,13^. Hematoxylin stains anionic regions like the nuclei of cells blue-purple, while eosin stains cationic regions like the cytoplasm and extracellular matrix pink^13^. This creates a contrast between the different components of the tissue, allowing pathologists to identify different structures and cells within the sample. H&E is the gold standard staining method used in pathology to visualize the tissue structure of biopsy samples. It is the primary means by which pathologists assess breast NCB samples to distinguish between malignant and benign breast tissue, as well as to determine the type and grade of cancer^14^.

The conventional tissue processing and staining procedures, despite being an essential part of histological analysis, are burdensome due to the significant costs, time, and expertise required^15^. However, advancements in label-free imaging technologies may have the potential to eliminate the need for these procedures, while preserving valuable biopsy samples for use in redundant or auxiliary screening procedures. Among the most promising ways of imaging tissue is an emerging technology called Photon Absorption Remote Sensing (PARS) microscopy. PARS enables simultaneous capture of contrast from both radiative and non-radiative relaxation processes following optical absorption, along with scattering contrasts in a tissue specimen^16^. The technique uses a picosecond-scale pulsed excitation laser to generate perturbations in the sample following absorption. The optical emissions from the radiative relaxation are broadly captured while the non-radiative contrast is measured as a percentage modulation in the back or forward scattering intensity of a secondary probe beam^16^. Depending on the excitation wavelength, PARS can provide sensitivity to a variety of chromophores including hemoglobin^17–19^, cytochromes^20,21^, DNA, collagen, elastin, and lipids^16,22,23^. Furthermore, by simultaneously capturing both absorption fractions, PARS provides additional contrast such as the quantum efficiency ratio (QER). QER is defined as the ratio of the non-radiative and radiative absorption portions (QER = P_nr_/P_r_) and is expected to yield additional biomolecular information^16^.

Recent works have employed an ultraviolet-based (UV, 266nm) whole slide imaging PARS system for label-free virtual histology^24–26^. Using the UV excitation source, PARS captures detailed nuclear contrast through the non-radiative relaxation of DNA absorption, as well as connective tissue contrast from the radiative relaxation of primarily collagen and elastin^27^. These contrasts are highly analogous to H&E staining and can be intelligently combined to virtually stain the sample. A deep learning-based image-to-image translation model is employed for H&E emulation and is trained on loosely registered PARS and H&E whole slide images pairs^26^. The resulting virtual H&E images demonstrate a high degree of structural and colour similarity; however, its diagnostic efficacy has not been thoroughly measured. To assess diagnostic equivalence, a concordance analysis can be used to quantify the level of agreement between the PARS virtual H&E images and the gold standard H&E stained samples.

Concordance rates refer to the degree of agreement between two or more pathologists who independently review the same tissue sample. Variability in the interpretation of breast core biopsies among pathologists can arise due to several contributing factors. These factors include the quality of the tissue sample obtained during the biopsy procedure and the level of experience of the pathologist. In addition, the amount of tissue available for histologic examination plays a role, wherein higher numbers of cores and longer cores tend to improve concordance among pathologists^28^. Therefore, in the context of breast NCBs, concordance rates are important to define because they reflect the diagnostic accuracy of the procedure, which typically informs subsequent surgical treatment decisions. In comparing PARS virtual H&E and true H&E, a high concordance rate would indicate a strong agreement, suggesting that the virtual histology method is successfully replicating the diagnostic information present in the traditional H&E staining. Evaluating concordance rates is crucial for validating PARS virtual H&E as a viable alternative to traditional H&E staining techniques in diagnostic applications.

We conducted a prospective study of seven independent breast tissue core biopsies representing a spectrum of known histologic findings spanning normal breast, ductal carcinoma *in situ*, invasive ductal carcinoma, and invasive lobular carcinoma. Unstained tissue was scanned with PARS microscopy to generate virtual H&E images, and then standard H&E staining of these same seven core biopsies was performed. The diagnostic characteristics of these images were assessed by multiple breast cancer expert pathologists, masked as to the origin of the images. We performed concordance analysis to define the diagnostic performances of the two imaging modalities, standard H&E, and PARS.

## 2. METHODS

### 2.1 Patient Materials

Tissues were acquired from the Cross-Cancer Institute (Edmonton, Alberta, Canada) through collaboration with clinical partners. The samples were obtained from anonymous patient donors, with all patient identifiers removed to ensure anonymity. The ethics committee waived the need for patient consent as these archival tissues were no longer necessary for patient diagnostics. Researchers were not provided with any information pertaining to the identity of the patients. Specimens were acquired under protocols (Protocol ID: HREBA.CC-18-0277) with the Research Ethics Board of Alberta and (Photoacoustic Remote Sensing (PARS) Microscopy of Surgical Resection, Needle Biopsy, and Pathology Specimens; Protocol ID: 40275) with the University of Waterloo Health Research Ethics Committee. All human tissue experiments were conducted in accordance with the government of Canada guidelines and regulations, including “Ethical Conduct for Research Involving Humans (TCPS2)”. The seven independent breast tissue core biopsies used in this study represented a spectrum of known histologic findings, spanning normal breast, ductal carcinoma *in situ* cancer, invasive ductal carcinoma, and invasive lobular carcinoma.

### 2.2 Sample Preparation

Core biopsy tissues underwent standard formalin-fixation, paraffin embedding, and sectioning, before being placed onto glass slides. Samples were fixed in formalin solution for a period of 24 to 48 hours, within 20 minutes of excision. Samples were then dehydrated with ethanol and treated with xylene to eliminate residual ethanol and fats. The samples were subsequently embedded in paraffin wax, creating formalin-fixed paraffin-embedded (FFPE) blocks. A microtome was then used to cut thin tissue sections (∼4-5μm) from the FFPE blocks. Tissue sections were placed on glass microscope slides and briefly baked at 60°C to evaporate excess paraffin.

### 2.3 PARS Microscope Imaging

Whole slide label-free PARS images were acquired from the unstained tissue sections using the PARS histology platform and architecture described in detail in^25^. In brief, the sample is precisely targeted with focused excitation pulses from a 50KHz 400ps 266nm UV laser (Wedge XF 266, RPMC). To achieve 40x imaging magnification, these excitation events are spaced 250nm apart, while three-axis mechanical stages move the sample across the objective lens in an “s”-shaped scanning pattern. At each excitation event, time-resolved radiative, non-radiative relaxation, and scattering signals are measured and compressed into single pixel intensity values. These intensity values collectively form the three co-registered label-free images.

To measure the radiative signal intensity, the spectrum of emitted photons is broadly collected with an avalanche photodiode (APD130A2, Thorlabs) and the peak amplitude value is recorded. To measure the non-radiative relaxation effect, time-domain photothermal and photoacoustic signals are recorded. This is done using a 405nm continuous-wave probe beam (OBIS-LS405, Coherent) which is coaxially aligned with the excitation spot. From this, a single non-radiative intensity value is extracted as the percentage modulation of the transmitted probe beam intensity before and after excitation. The scattering intensity of the sample is determined by calculating the average probe transmission intensity prior to excitation. Both the excitation and detection beams are focused onto the sample using a 0.42 numerical aperture (NA) UV objective lens (NPAL-50-UV-YSTF, OptoSigma). The transmitted probe light and radiative photons are collected using a 0.7NA objective lens (278-806-3, Mitutoyo). The radiative spectrum (>266nm) and 405nm detection wavelength are then spectrally separated prior to measurement.

The entire sample is scanned in 500x500μm parts which are later stitched back together into a single whole slide image. The 405nm scattering contrast is primarily used to find and maintain sharp focus across the sample while the radiative and non-radiative images are primarily used for virtual staining.

### 2.4 H&E Staining and Digital Image Acquisition

After all samples were imaged with the PARS microscope, standard H&E staining was performed on each of the seven core biopsies. Digital whole slide H&E images were then acquired at 40x resolution using a standard brightfield microscope (Morpholens 1, Morphle Digital Pathology).

### 2.5 PARS Virtual H&E Colourization

A cycle-consistent generative adversarial network (CycleGAN), first developed by Zhu et. al^29^, was employed to convert the PARS label-free data to virtual H&E images. This deep-learning based image-to-image translation model has previously been used for virtual H&E staining of PARS label-free contrast^26^. Here, with the exception of the Noise2Void denoising algorithm, the same training workflow and data preparation process was used, and a virtual H&E model was trained using a distinct set of whole slide image pairs. These additional training samples underwent the same tissue processing, imaging, and staining procedures as the core needle biopsies.

In brief, the PARS label-free whole slide images are first loosely registered to their corresponding ground truth H&E pairs using a simple affine transform with three registration points. Images are then cut into 512x512px (128x128um) tile pairs for use in model training.

Prior to slicing, the PARS label-free contrasts are combined into a single total absorption (TA) coloured image where the radiative channel is blue, and the non-radiative channel is red. An example TA whole slide image and corresponding training pairs can be seen in Figure 1 alongside its corresponding ground truth H&E image. The virtual staining model in this study was trained on roughly 1000 training pairs. Once the virtual staining model is trained, the same model was then applied to all seven breast core needle biopsies, forming the virtual and real H&E pairs.

**Figure 1:**
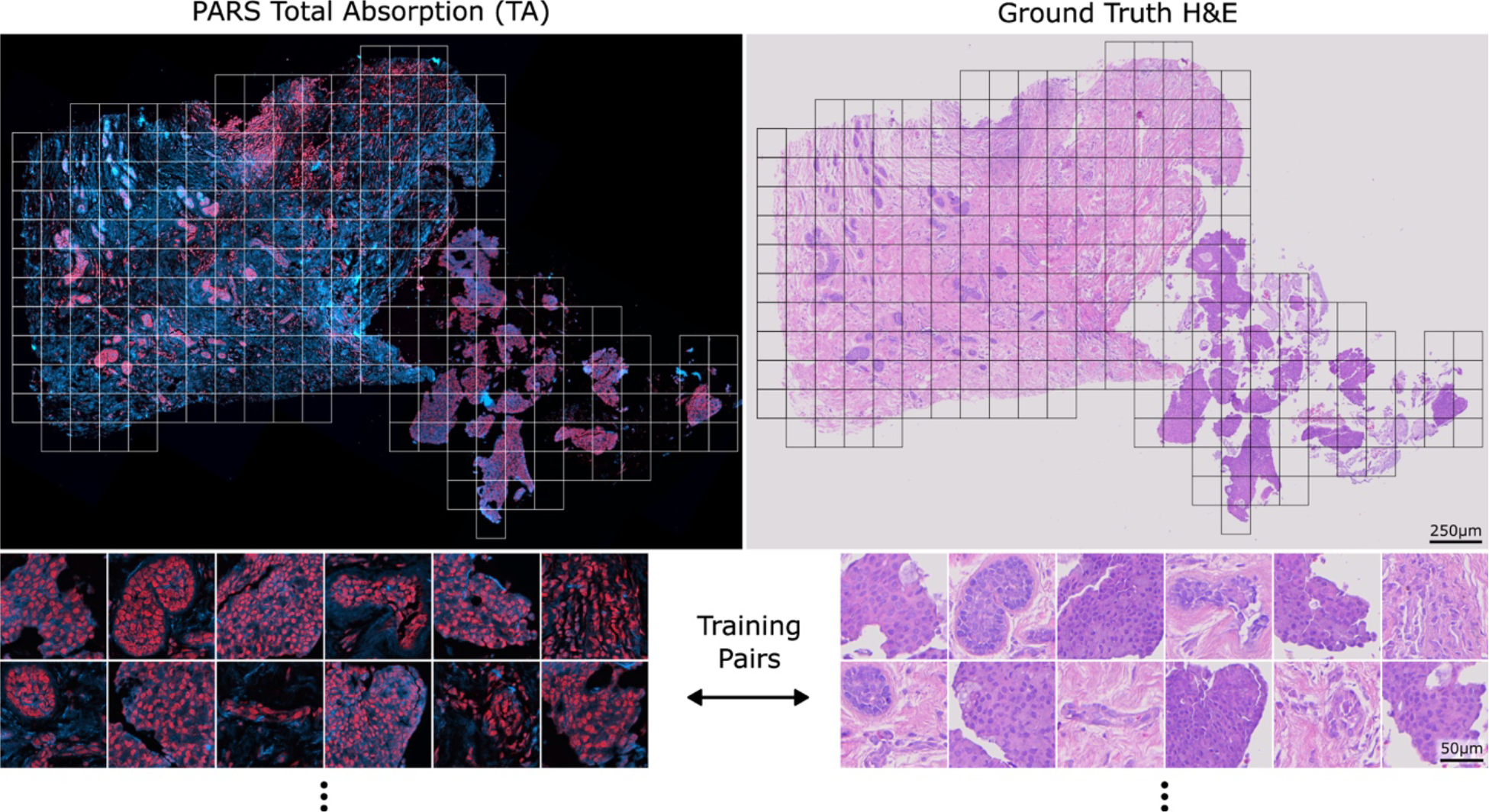
An example PARS total absorption (TA) image side-by-side with its corresponding ground truth H&E whole slide image. Example 512x512px pairs that can be used to train the model can be seen below each image.

### 2.6 Evaluation by Expert Pathologists

The PARS virtual H&E and true H&E images were randomly oriented and displayed in a pre-specified, custom random order generation algorithm designed to maximize the distance between the two image pairs (PARS and true H&E) for each individual sample. Each of the 14 images were placed on a customized web-based histology visualizing software platform without any identification except core biopsy #1 through core biopsy #14. The order of sample display was P2, P5, T1, T4, T3, T6, T2, P1, P7, P4, T5, T7, P3, and P6 where ‘P’ corresponds to PARS virtual H&E and ‘T’ corresponds to true H&E. Each of 14 images were provided independently to breast cancer focused board-certified anatomic pathologists, and 7 surveys were completed. The pathologists were masked to the clinicopathologic details of the cases and the origin of the digital images (either True H&E or PARS virtual H&E). Each pathologist was asked to score each image on the parameters shown in Table 1, including histologic diagnosis, grade of *in situ* disease, grade of invasive disease, and the origin of the digital image (Table 1).

**Table 1:** Survey questionnaire given to pathologists for each of the 14 total images.

|  |  |  |  |  |  |
| --- | --- | --- | --- | --- | --- |
| **1. The primary tissue diagnosis is:** |  |  |  |  |  |
|  | Invasive ductal carcinoma | Invasive lobular carcinoma | DCIS | Normal glandular tissue | Image inadequate for diagnosis |
|  | ☐ | ☐ | ☐ | ☐ | ☐ |
| **2. DCIS necrotic score:** |  |  |  |  |  |
|  | No In Situ disease present | Grade 1 | Grade 2 | Grade 3 | Not assessable |
|  | ☐ | ☐ | ☐ | ☐ | ☐ |
| **3. DCIS nuclear grade:** |  |  |  |  |  |
|  | No In Situ disease present | Grade 1 | Grade 2 | Grade 3 | Not assessable |
|  | ☐ | ☐ | ☐ | ☐ | ☐ |
| **4. Evaluation for invasive disease:** |  |  |  |  |  |
|  | No invasive disease | Grade 1 | Grade 2 | Grade 3 | Not assessable |
| Architectural | ☐ | ☐ | ☐ | ☐ | ☐ |
| Nuclear | ☐ | ☐ | ☐ | ☐ | ☐ |
| Mitotic | ☐ | ☐ | ☐ | ☐ | ☐ |
| **5. Type of image: Is this image from FFPE H&E stained tissue?** |  |  |  |  |  |
|  | Yes, this is H&E | No, this is not H&E | Uncertain |  |  |
|  | ☐ | ☐ | ☐ |  |  |

### 2.7 Statistical Analysis

Concordance analysis is a statistical method used to measure the agreement between two or more raters or observers in their interpretation or classification of a set of data. Concordance may be measured through the Cohen’s and Fleiss’ kappa coefficients. Cohen’s kappa coefficient is a measure of inter-rater reliability that takes into account the possibility of agreement occurring by chance^30^. It is used to determine whether two raters agree beyond what would be expected by chance alone. Fleiss’ kappa is an extension of Cohen’s kappa to compare more than two raters^31^.

Kappa values range from -1 to 1, with a value of 1 indicating perfect agreement and a value of 0 indicating agreement no better than chance. Negative values indicate agreement worse than chance. Interpretation of kappa values vary, but a value in excess of 0.6 is considered “substantial”^32^ or “good”^33^.

This method has several advantages over other measures of agreement, including its ability to account for chance agreement and its robustness to variations in the prevalence of different categories of data^30^. All calculations were performed in R statistical software^34^.

## 3. RESULTS

### 3.1 Example Whole Slide Image Pairs

Figures 2 and 3 show two exemplary sets of PARS virtual H&E and real H&E images employed in this study. At the top of each figure is the raw total absorption (TA) PARS image serving as the input to the virtual staining algorithm. Both of these figures show examples of invasive ductal carcinomas, with higher magnification regions showcasing irregular malignant glandular structures infiltrating a fibrofatty stroma, characteristic of invasive breast carcinoma. One benefit to the virtual H&E stains is that they all share consistent stain colouring, which matches the colouring of the training dataset. In contrast, staining colours for true H&E images can vary depending on specifics of the preparation, digitization and storage of the tissue samples^35^. As such, the virtual H&E images in figures 2 and 3 share similar staining colours whereas their true H&E counterparts exhibit a slight difference in colours. Nonetheless, both the virtual and H&E images achieve excellent epithelial and stromal contrast and highlight the same tissue structures.

**Figure 2:**
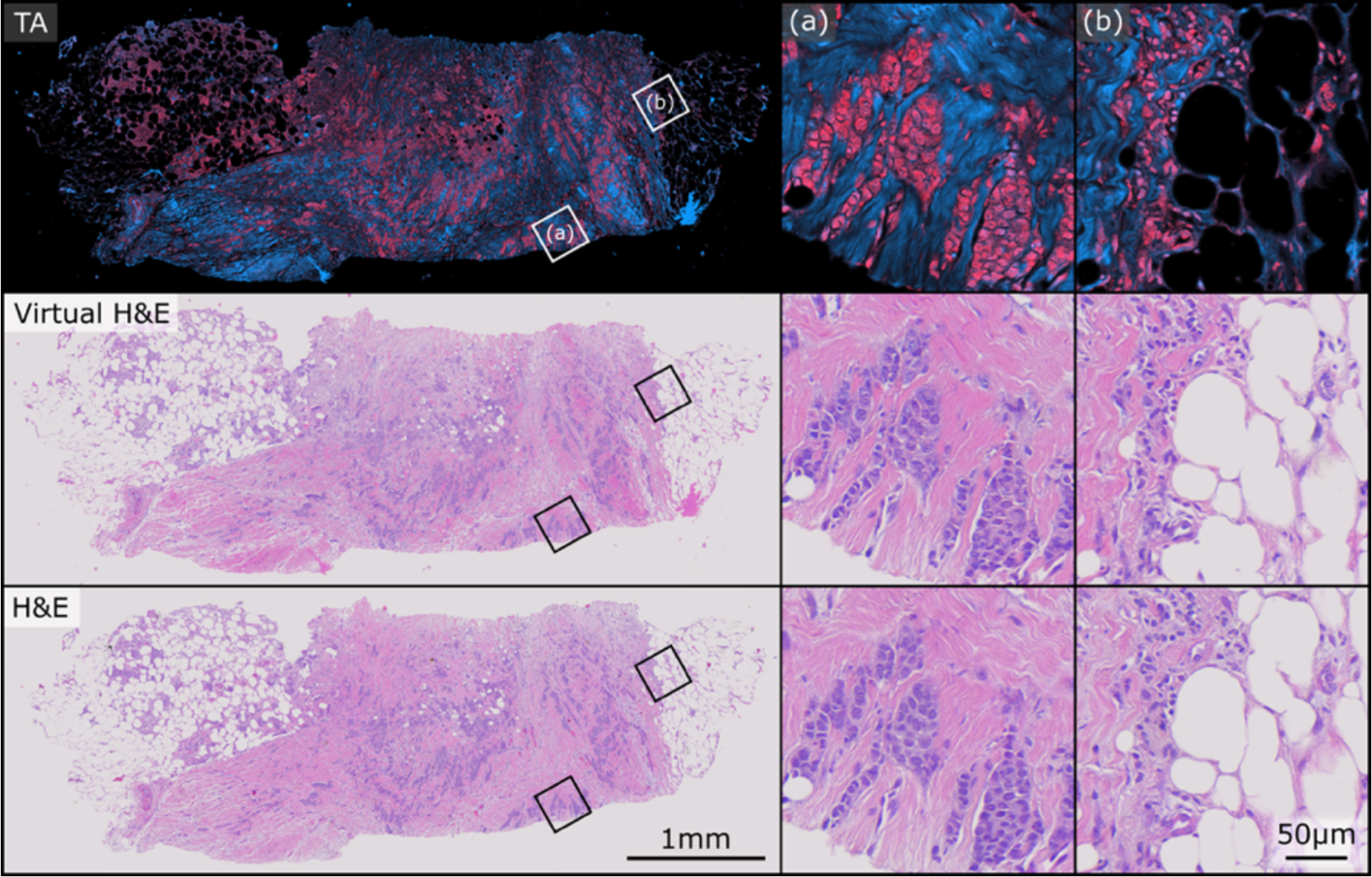
Example PARS total absorption (TA), virtual H&E, and true H&E pair used in this study. The sample exhibits closely matched staining colours between the virtual and real representations. (a) and (b) depict two regions of higher magnification on the sample

**Figure 3:**
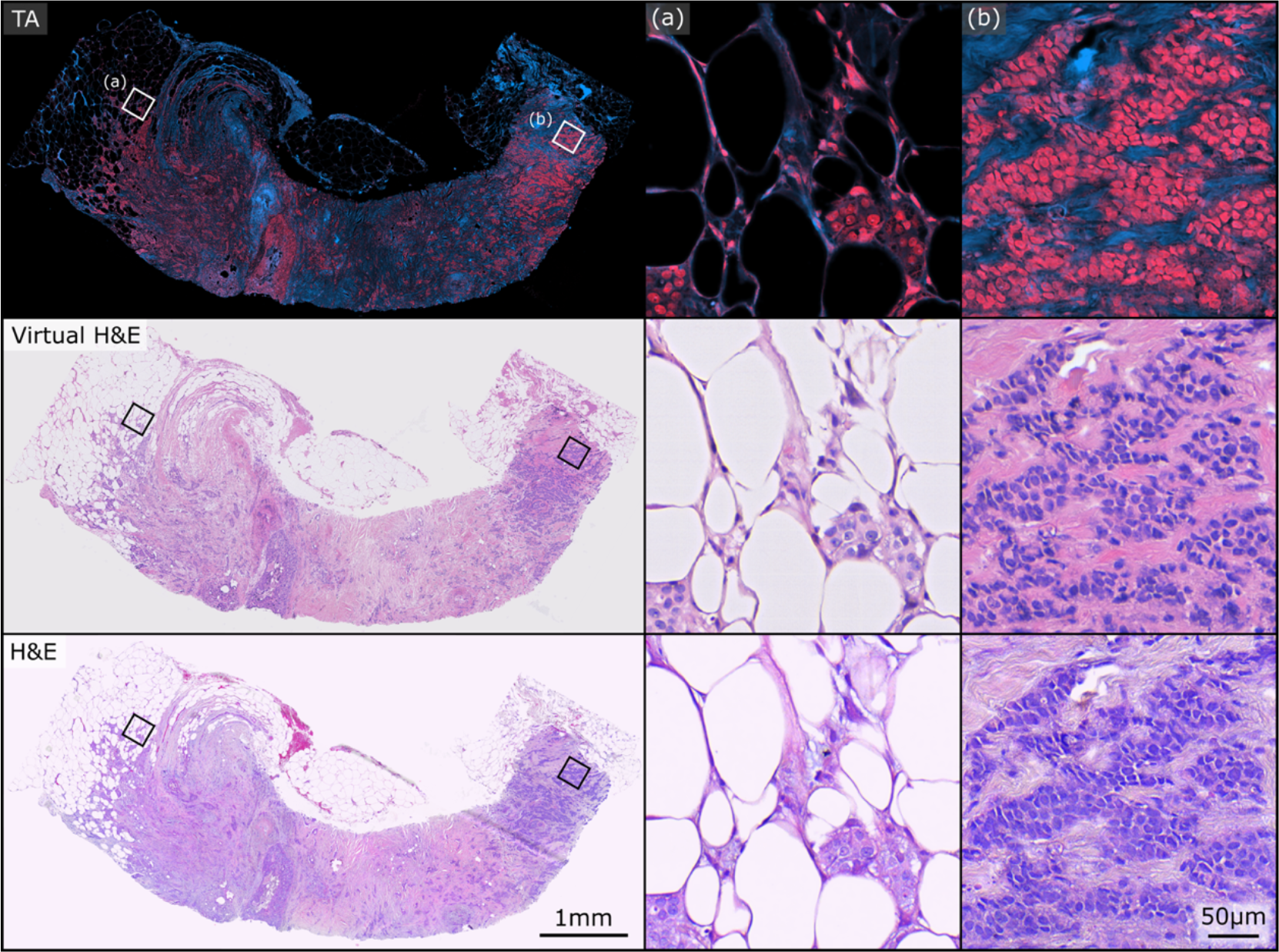
Example PARS total absorption (TA), virtual H&E, and true H&E pair used in this study. Both the virtual and real H&E images exhibit excellent epithelial and stromal contrast and highlight the same tissue structures. (a) and (b) depict two regions of higher magnification on the sample

### 3.2 Image Origin

For each image, either true H&E or PARS virtual H&E, respondents were asked to identify the origin of each image, whether it was a true FFPE H&E stained slide (yes), not a true FFPE H&E stained slide (no), or uncertain. Three raters responded ‘Yes’ when asked if a PARS image was a true H&E image for all seven images. The fourth respondent reported ‘Uncertain’ for all seven PARS and H&E images. For the remaining three pathologists, PARS images were misidentified as true H&E images 0/7, 1/7, 3/6 times and true H&E images were misidentified as virtual H&E images 1/7, 3/7, 3/6 times (the final pathologist only responded to this question for six image pairs). These results show that masked pathologists were unable to reliably distinguish between conventional H&E and PARS virtual H&E.

### 3.3 Primary Diagnosis

Respondents were asked to make a primary diagnosis for both the PARS virtual H&E images as well as the true H&E images. All respondents were able to make a primary diagnosis for each whole slide image with the exception of the fourth respondent who selected ‘Image inadequate for diagnosis’ precisely once.

If all primary diagnosis responses are combined into either a high level ‘cancer’ or ‘benign’ category, out of the 48 image-pair assessments (excluding one diagnosis of ‘image inadequate’), there was only one disagreement between an H&E and PARS pair (kappa = 0.921). In total, 40 image-pairs were both assessed as cancerous, while the remaining 7 image-pairs were both assessed as benign. This indicates there was reliable discrimination between cancerous and benign cases. Here ‘cancer’ comprises the diagnosis of invasive lobular carcinoma, invasive ductal carcinoma, and DCIS.

For the specific cancer subtypes, a concordance analysis for the primary diagnosis among the seven pathologists was done for both true H&E only and PARS virtual H&E only. The Fleiss’ kappa for agreement between rater for true H&E images was 0.639. The Fleiss’ kappa for agreement between rater for PARS virtual H&E images was 0.620. Next, a pairwise comparison was conducted, calculating Cohen’s kappa, to assess the concordance of the primary diagnosis between the PARS virtual H&E and true H&E images. Four of the seven pathologists (raters 3, 5, 6, 7) agreed on the primary diagnosis for all seven image pairs (kappa = 1). The first respondent disagreed on the primary diagnosis for a single image pair (kappa = 0.611). The second and fourth respondents disagreed on the primary diagnosis for two image pairs (rater 2, kappa = 0.364; rater 4, kappa = 0.417). Table 2 shows a comparison between the primary diagnosis given to the H&E and PARS image pairs.

**Table 2:** Summary of pathologist responses to question “the primary tissue diagnosis is”.

| PARS Diagnosis | H&E Diagnosis |  |  |  |  | Total |
| --- | --- | --- | --- | --- | --- | --- |
|  | IDC | ILC | DCIS | Benign | Image Inadequate |  |
| IDC | 36 | 0 | 1 | 1 | 0 | 38 |
| ILC | 1 | 1 | 0 | 0 | 0 | 2 |
| DCIS | 1 | 0 | 0 | 0 | 0 | 1 |
| Normal Tissue | 0 | 0 | 0 | 7 | 0 | 7 |
| Image Inadequate | 1 | 0 | 0 | 0 | 0 | 1 |
| Total | 39 | 1 | 1 | 8 | 0 | 49 |

*Note:* IDC, invasive ductal carcinoma; ILC, invasive lobular carcinoma; DCIS, ductal carcinoma *in situ*

### 3.4 Evaluation of Tissue Gradings

A similar analysis to the primary diagnosis response was done to assess the concordance for the evaluation of invasive tissue components, namely: architectural score, nuclear grade, and mitotic index. In Table 3, a Fleiss’ kappa coefficient was computed for the within-H&E only invasiveness gradings and the within-PARS only invasiveness gradings. This was done to first observe the concordance among pathologists for H&E only and for PARS only and contrast it with the concordance result for a pairwise comparison of concordance between H&E and PARS. For the pairwise comparison, a Cohen’s kappa coefficient was computed for each rater and the average coefficient is reported in Table 3. The kappa coefficients were computed from responses of all image pairs excluding image pair six. Image pair six was not involved because it was given the primary diagnosis of ‘normal glandular tissue’ from all raters except one (rater 2, PARS image).

**Table 3:** Inter-rater reliability of invasive cancer grading.

| Evaluation Component | Comparison | Kappa Coefficient** |
| --- | --- | --- |
| Invasive Architectural Score | Within - H&E | 0.553 |
|  | Within - PARS | 0.600* |
|  | H&E - PARS | 0.420 |
| Invasive Nuclear Grade | Within - H&E | 0.051 |
|  | Within - PARS | 0.058 |
|  | H&E - PARS | 0.188 |
| Invasive Mitotic Index | Within - H&E | 0.125 |
|  | Within - PARS | 0.148 |
|  | H&E - PARS | 0.032 |

\* Corresponds to 'substantial' agreement.
\*\* The pairwise H&E - PARS comparison are reported as the mean Cohen's kappa coefficient among raters.

In some cases, for both true H&E and PARS images, pathologists were unable to assess or submit a grading between 1-3. Table 4 summarizes the total number of ‘not assessable’ responses for each image type across all 49 image pair assessments. ‘Both’ is the number where the rater categorized both H&E and PARS images in the same pair as not assessable for that evaluation.

**Table 4:** Distribution of “not assessable” responses.

|  | DCIS<br>Necrotic | DCIS<br>Nuclear | Invasive<br>Architectural | Invasive<br>Nuclear | Invasive<br>Mitotic |
| --- | --- | --- | --- | --- | --- |
| H&E | 1 | 7 | 1 | 5 | 15 |
| PARS | 2 | 4 | 1 | 6 | 14 |
| Both | 1 | 3 | 0 | 3 | 8 |

For each rater, there was widespread agreement between which images were assessable or not. The accessibility was highest for the architectural score (48/49 H&E, 48/49 PARS), slightly lower for nuclear grade (44/49 H&E, 43/49 PARS), and much lower for the mitotic index (34/49 H&E, 35/49 PARS). Additionally, the number of image pairs where pathologists agreed that both PARS and H&E was either assessable or not assessable followed the same trend. Agreement was observed in 47/49 image pairs for the architectural score, 44/49 for nuclear grade, and 36/49 for the mitotic index.

## 4. DISCUSSION

### 4.1 Study Summary and Key Findings

In this pilot validation study, seven pairs of PARS virtual and conventional H&E images were assessed by seven pathologists masked with respect to the origin of the images. Comparative analysis of PARS and H&E using the standardized synoptic reporting of the single core biopsy images demonstrated several key findings. Both PARS virtual and conventional H&E images were of diagnostic quality, and reliably allowed the discrimination of cancer (the aggregate diagnoses of invasive ductal carcinoma, invasive lobular carcinoma, and DCIS) from benign breast tissue. With respect to the primary diagnostic categories above, the raters show almost identical agreement across H&E images as they do across PARS images. Furthermore, the diagnoses made by our pathologists viewing conventional H&E were comparable to the diagnoses of pathologists viewing PARS virtual H&E; with the granular categorization of each image into the five primary diagnostic results, overall concordance among pathologists was substantial (kappa > 0.6) and was not meaningfully higher with conventional H&E images rather than the PARS virtual H&E images. Four of the seven pathologists agreed on the primary diagnosis for all seven image pairs, one pathologist disagreed on the primary diagnosis for a single image pair, and two pathologists disagreed on the primary diagnosis for two image pairs.

### 4.2 Interpretation of Findings in Context

Diagnostic reproducibility in breast cancer histology remains suboptimal and underpins the difficult of evaluating new histologic techniques. Within the context of our study, PARS-based breast core biopsy imaging was equivalent to more traditional digital histopathology using H&E stained slides. Inter-observer discordance appeared lower than previously reported intra-observer discordance. For example, pathologists given the same breast cancer biopsy material on which they had previously issued a diagnostic report, separated by a six month interval, exhibit surprisingly low intra-observer agreement rates of 92% for invasive breast cancer, 84% for DCIS, and 53% for benign with atypia^36^. Similarly, pathology review of the original breast cancer needle core biopsy in a pre-operative quality assurance process identified 403 discordance cases out of 4950 (∼8%)^37^. Furthermore, histologic interpretation and grading of core needle biopsies is dependent on the quantity of available tissue, which in our study was limited to a single core biopsy per case. The authors could find no published literature on the inter-rater reliability of breast cancer core biopsy gradings, so it is difficult to put the grading data we generated in context, other than to state that grading concordance was poor (0.6 or less) with both conventional and PARS virtual H&E.

### 4.3 Strengths, Limitations and Future Research

Among the strengths of our study was the use of masked pathologists, the use of 40x digital scanning equivalent to standard digitized conventional H&E images, and efforts to reduce confounding observer recognition of serially presented images by re-orienting pairs of biopsies and maximizing their sequence separations. Furthermore, our study used the identical tissue for the two images, rather than adjacent slides, allowing direct cell to cell concordance of the images. Limitations of our study include its relatively small sample size, and the selection of tissues representing only the most common histologic findings on breast biopsy. Future research should aim to conduct larger, studies with additional samples to substantiate and build upon our findings. Furthermore, these tissues were imaged in late 2022, and these results may underestimate the true diagnostic utility of PARS compared to conventional H&E given recent and meaningful improvements in PARS imaging techniques ^26,38^. This includes the addition of a 532nm (green) excitation source with its own non-radiative and radiative contrasts, as well as multichannel PARS non-radiative signal extraction ^38^ and image denoising ^26^. Consequently, using our current state-of-the-art PARS histology might further improve these results.

### 4.4 Implications and Conclusions

This prospective cohort study provides evidence supporting the effectiveness of PARS microscopy for the diagnostic interpretation of human breast tissue core biopsies. The images were deemed of diagnostic quality by expert breast cancer pathologists. The key consideration of cancer vs. benign tissue was reliably distinguished in both conventional and PARS virtual H&E histology images. Similarly, cancer subtypes were reliably distinguished with both techniques.

While the initial diagnosis of breast cancer is typically made by conventional H&E evaluation of core biopsies, the complexity of the tissue preparation and staining frequently requires one week or more before the pathology report is available. PARS is an imaging technique that can be applied not only to fixed, unstained tissues, as in this specific study, but also to freshly resected specimens and in vivo examination of tissue. As such, PARS has the potential to dramatically reduce diagnostic timelines.

Finally, PARS is a non-destructive process that generates a rich dataset suitable for analysis by artificial intelligence algorithms, which are being successfully applied to cancer diagnosis of digital histology^39,40^. The virtual colourization process already leverages in-house developed AI algorithms, and analysis by AI would be a natural extension of this process. The unstained tissue remains suitable for any additional subsequent analyses, which allows downstream standard-of-care processing of samples to be unaffected. Consequently, PARS virtual histology has the potential to both improve the speed and the accuracy of diagnostic interpretation of breast histology, reduces the consumption of limited biopsy tissue, and is, in principle, widely applicable to histologic evaluation of benign and malignant tissues of any origin. Moreover, this study was limited to human breast cancers but should be directly applicable to all other types of biopsied organs since tissue preservation and H&E staining are the same procedure for all types of tissues.

## Data Availability

All data produced in the present study are available upon reasonable request to the authors.

## Acknowledgments and Funding Sources

The authors would like to thank the Cross-Cancer Institute in Edmonton, Alberta for providing human breast tissue samples. The authors would also like to thank Dr. Ellen Cai, Dr. Julinor Bacani, Dr. Judith Hugh, Dr. Murray Savard, Dr. Richard Berendt, Dr. Erene Farag, and Dr. Nadia Giannakopoulos for participating in the survey and questionnaire. The authors thank the following sources for funding used during this project. Natural Sciences and Engineering Research Council of Canada (DGECR-2019-00143, RGPIN2019-06134); Canada Foundation for Innovation (JELF #38000); Mitacs Accelerate (IT13594); University of Waterloo Startup funds; Centre for Bioengineering and Biotechnology (CBB Seed fund); illumiSonics Inc (SRA #083181); New frontiers in research fund – exploration (NFRFE-2019-01012); The Canadian Institutes of Health Research (CIHR PJT 185984).

## Author Contribution Statement

JEDT processed the PARS images for model training, applied the virtual H&E staining, and organized the images on the custom web-based histology visualizing software platform. JEDT and BRE built the PARS optical system and PARS imaged all the breast needle core biopsy tissue sections. HG performed all tissue processing and H&E staining procedures as well as digitally scanned the true H&E images. JRM, DD, and GB designed the survey questionnaire and provided clinical feedback and consultation in the assessment of the results. JRM launched the final survey and recruited pathologists for the questionnaire. MPW performed the statistical analysis of the results and assisted in writing the results section of the manuscript. JEDT and JRM were the major contributors in writing the manuscript. All authors read, provided edits, and approved the final manuscript. PHR directed and organized the project and oversaw experimental work and manuscript writing as the principal investigator.

## Data Availability Statement

Datasets used and analysed during this study are available from the corresponding author on reasonable request.

